# A Data-Informed Approach for Analysis, Validation, and Identification of COVID-19 Models

**DOI:** 10.1101/2020.10.03.20206250

**Authors:** S. Yagiz Olmez, Jameson Mori, Erik Miehling, Tamer Başar, Rebecca L. Smith, Matthew West, Prashant G. Mehta

## Abstract

The COVID-19 pandemic has generated an enormous amount of data, providing a unique opportunity for modeling and analysis. In this paper, we present a data-informed approach for building stochastic compartmental models that is grounded in the Markovian processes underlying these models. Our initial data analyses reveal that the SIRD model – susceptiple (S), infected (I), recovered (R), and death (D) – is not consistent with the data. In particular, the transition times expressed in the dataset do not obey exponential distributions, implying that there exist unmodeled (hidden) states. We make use of the available epidemiological data to inform the location of these hidden states, allowing us to develop an augmented compartmental model which includes states for hospitalization (H) and end of infectious viral shedding (V). Using the proposed model, we characterize delay distributions analytically and match model parameters to empirical quantities in the data to obtain a good model fit. Insights from an epidemiological perspective are presented, as well as their implications for mitigation and control strategies.

## I. Introduction

The COVID-19 pandemic has infected individuals in nearly every country on earth. The pervasive spread of the underlying virus, SARS-CoV-2, combined with modern day data collection practices, has resulted in vast amounts of epidemiological data. This has provided researchers with a unique opportunity to not only model, analyze, and mitigate COVID-19, but also to develop best practices to quickly respond to future pandemics.

From a modeling perspective, compartmental models are the predominant approach for describing the progression of diseases. These models describe an aggregated view of disease dynamics by partitioning a given population into groups, termed *compartments*, and studying how the size of the groups evolve in time, typically via ordinary differential equations (ODEs). In the case of modeling COVID-19, some simple models, such as the SIR model with three compartments (states) – susceptible (*S*), infected (*I*), and recovered (*R*) – have been studied [1]–[3]. Such models have generated many insights, proving to be useful in the estimation of model parameters (e.g., estimation of the basic reproduction number *R*_0_), prediction of the future evolution of the disease (e.g., the predicted number of infected agents), and the analysis and design of mitigation and control strategies (e.g., the effects of lockdown or mask wearing, tradeoffs between epidemiological and optimal costs).

Since the beginning of the pandemic, several extensions of the basic SIR model have been considered to include additional states such as exposed (*E*), asymptomatic (*A*), quarantined (*Q*), hospitalized (*H*), and death (*D*) [1]–[10]. For example, the SIDARTHE model also considers a patient’s health and testing status [10]. Although ODE models remain the most popular, stochastic extensions of these models have also been considered to better capture the effect of un-modeled noise [11]–[15]. Other types of modeling options include neural networks and artificial intelligence [16], autoregressive integrated moving average (ARIMA) [17], and agent-based approaches [18], [19].

Although the SIR ODE model (and its many deterministic and stochastic generalizations) is useful, these models represent (mean-field) approximations of the underlying dynamics. The epidemiological states are more fundamentally described for a single agent who is first susceptible and then becomes infected and eventually recovers. The main assumption underlying the mean-field approximation leading to an ODE model is that the transitions between the epidemiological states are Markovian. In the case of COVID-19, a state transition diagram (see Fig. 1) is useful to visualize the evolution of the disease for a single agent. For the *S* →*E* transition, one typically assumes that the rate parameter scales proportionally with the number of infected agents in the population. In the language of interacting particle systems, the *S* →*E* transition depends upon the interaction amongst agents. In contrast, the transition when a susceptible agent is first exposed to (or infected by) the virus depends upon the population. The remaining transitions, however, are not affected by the population, depending instead solely upon the intrinsic characteristics of the agent (e.g. age, gender, pre-existing conditions).^1^

**Fig. 1:**
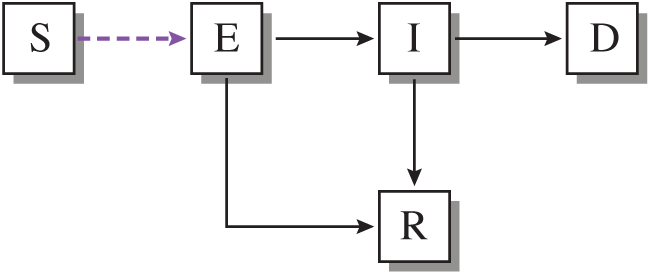
An intuitive compartmental model for COVID-19. The (purple/dashed) transition from susceptible (*S*) to exposed (*E*) depends on the population. The remaining (black) transitions depend upon characteristics of individual agents. Exposed and infected (*E*) can lead directly to recovery (*D*) or recovery (*R*). Note that all transitions are directed. (*R*), or to infected and symptomatic (*I*). (*I*) can then lead to either death

We have two goals in this paper. The first goal is to examine the Markovian assumption underlying the agent-dependent transitons (e.g., *I* →*R, I* →*D*). This is done through collecting and analyzing data related to observed transition time delay distributions for a large number of infected agents. The second goal is to develop and apply a data-informed methodology to construct Markov models with hidden states that better fit the delay distribution and fatality rate data. The approach is demonstrated for real data collected as part of the first goal.

The proposed data-informed approach allows for an incremental construction of compartments such that the model sufficiently explains the data, while being based on epidemiological evidence for SARS-CoV-2. In contrast with the current body of research, this work serves to highlight the importance of data to disease modeling for COVID-19, where models are built from the insights provided by the data. This model building process is demonstrated through the progression from a model of low to high complexity based on transition distribution fitting.

The outline of the remainder of this paper is as follows. In Sec. II, the epidemiological data used in this study is briefly summarized. The main aspects of analysis and model construction appear in Sec. III. A discussion of the resulting epidemiological insights and its implications for control appear in Sec. IV.

## II. Epidemiological Data

Much of the publicly-available COVID-19 data is aggregate data, such as the number of cumulative and daily new infections, death count, testing data, etc. These data are not directly useful for the type of epidemiological modeling considered here. For our purposes, the so called *anonymized line-list data*^2^ which charts the progression of disease in anonymized agents are needed. Such data require prior IRB approval, which is currently pending for our project. In the interim, we have gathered all the publicly-available data on delay distributions between various transitions of interest. The nomenclature for the epidemiological states appears in Table I. A summary of data appears in Table II. It is noted that no data was found that dealt with the transition from susceptible state (*S*). The data for exposure (*E*) were highly uncertain. Consequently, the remainder of this paper focuses on remaining transitions {*I, H, R, D*}.

**TABLE I:** Nomenclature for the Epidemiological States

| Symbol | Epidemiological Meaning |
| --- | --- |
| $S$ | susceptible |
| $E$ | exposure |
| $I$ | symptomatic infection |
| $H$ | hospital admission |
| $R$ | recovered |
| $D$ | death |

### Summary of data

The preliminary epidemiological data upon which our analysis is based consist of a collection of 10 de-identified, open-source datasets acquired online [20]–[28]. The countries of origin for these data are United States, China, and South Korea, as well as one dataset of international travelers, which encompasses individuals from 27 nations. All datasets contain key demographic information regarding patient age and sex, and some include additional information about comorbidities and symptoms. In order to examine the impacts of age on COVID-19 infection outcomes, which have been seen in clinical settings [29]– [31], we have stratified the data by ages 0-19, 20-59, and 60+ when possible.

We summarize next the data on delay distributions relevant to the following transitions important to COVID-19 modeling:

#### (i) Delay distribution for *I* →*H* transition

The NIH data, as listed in line 2 of Table II, contain data for 814 individuals. However the individuals with non-positive delays are removed from the dataset. This reduces the number to 371. Fig. 2a depict the histogram of the ages in the dataset. Fig. 3b and 3c depict the time delay distributions for adults and elderly together with the maximum likelihood exponential and gamma distribution fits. In contrast to *I* → *R* and *I* →*D* datasets, the delay distribution for *I* →*H* transition is reasonably approximated by the exponential distribution for both adults and elderly.

**TABLE II:**
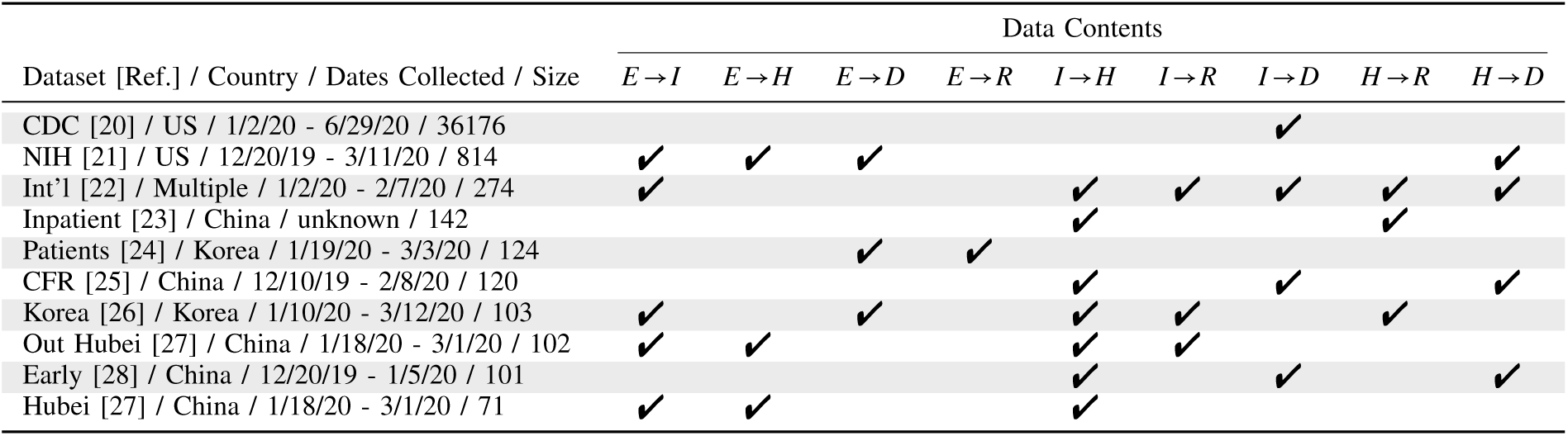
Breakdown of datasets and their contents.

**Fig. 2:**
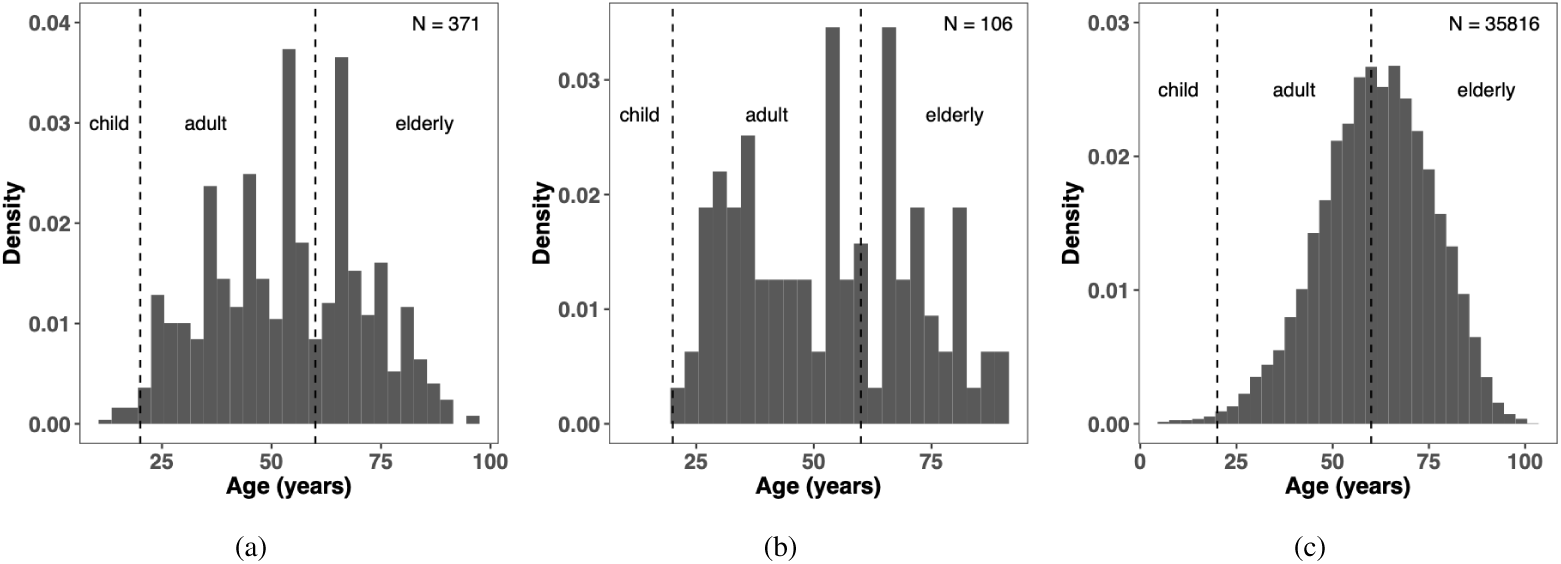
Histogram of ages in (a) *I* → *H*, (b) *I* → *R*, (c) *I* → *D* datasets. Individuals with non-positive delays are removed from the datasets.

**Fig. 3:**
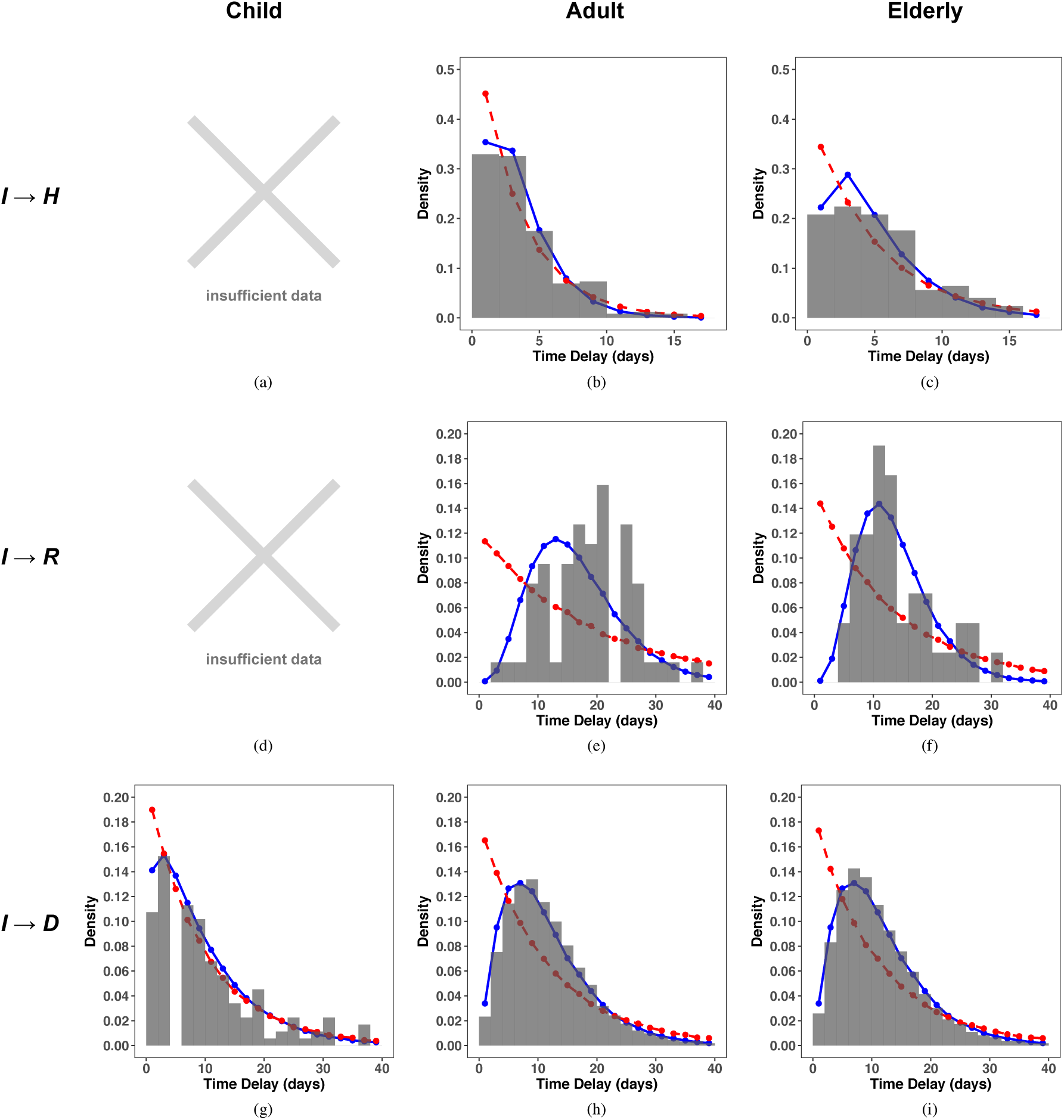
Time delay distributions for three age categories. Maximum likelihood gamma fits are marked blue and exponential fits are marked red. There were insufficient data to plot the *I* →*H* and *I* →*R* transitions for the child class (as seen in (a) and (d)).

#### (ii) Delay distribution for *I R* transition

Compared to *I* → *H* and *I* → *D* transitions, there is much less publicly available data for the *I* →*R* transition. For this transition, data from three different datasets were consolidated: data from China with 56 datapoints, data from Korea with 21 datapoints, and a data from international passengers with 29 datapoints. On account of the small number of datapoints, three datasets are combined to carry out the analysis. Fig. 2b depicts the histogram of ages in the combined dataset. Fig. 3e and 3f depict the time delay distributions for the different age groups together with the maximum likelihood exponential and gamma distribution fits. As also observed for the *I* → *D* transition, these distributions are not exponential and moreover distributions for adults and elderly are somewhat similar (although the latter point is difficult to conclude based on the small number of data points in these datasets).

#### (iii) Delay distribution for *I* →*D* transition

CDC data, as listed in line 1 of Table II, are the most comprehensive source for delay distribution of the *I* →*D* transition. The CDC dataset contains data for 36,176 individuals including age and sex; 35,816 of which have positive delay values. Fig. 2c depicts the histogram of ages in the CDC dataset. Fig. 3g-3i depict the time delay distributions for three different age categories – children, adults, and elderly – along with the maximum likelihood exponential and gamma distribution fits. The data and the fits allow us to make the following observations:

i. The delay distributions are clearly not exponential for adults and the elderly.
ii. The distributions for adults and the elderly (two age categories that dominate the dataset) are remarkably close to each other. This is despite significant differences in case fatality rates.

In the following section, our aim is to fit compartment models based on using these delay distributions.

## III. Data Analyses

This section introduces a process of fitting a continuous time Markov chain model to the data. This is achieved by augmenting the chain one state at a time with the goal of matching empirical quantities of the data to their theoretical values expressed in terms of rate parameters. As will be shown, we obtain a model that describes the data well.

### A. Notation and a Basic Model

We begin by introducing the notation used throughout our analyses as well as some useful results. For the Markov chain depicted in Fig. 4a, the observed quantities that appear in the data correspond to the following set of random variables:

**Fig. 4:**
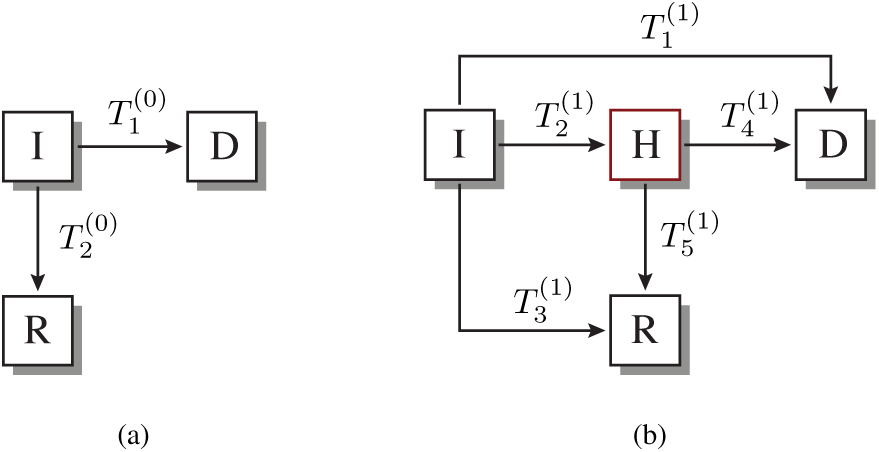
(a) No hidden states, 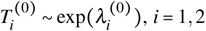. (b) One hidden state, 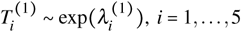

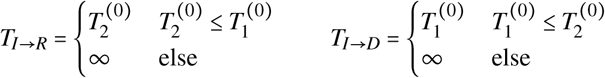

The distributions for these random variables are stated in the following elementary proposition.

#### Proposition 1.

*Consider the Markov chain depicted in Fig. 4a. Then, conditioned on the respective events* {*T*_*I*→*R*_ < ∞} *for T*_*I*→*R*_ *and* {*T*_*I*→*D*_ < ∞} *for T*_*I*→*D*_,

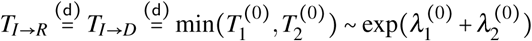

*where* 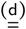 *denotes equivalence in distribution*.

The above proposition is useful in examining the model’s validity. In fact, the model is *not* valid due to the following:

i. The observed distributions for *T*_*I*→*R*_ and *T*_*I*→*D*_ are not exponential; and
ii. The observed distributions for *T*_*I*→*R*_ and *T*_*I*→*D*_ are not identical.

The goal of the remainder of this section is to consider enhancements of the basic model, via the addition of hidden states, to better fit the data.

### B. One Hidden State

The simplest possible enhancement of the basic model is to introduce one hidden state. The states under this augmented model are {*I, H, R, D*}, where *I, R, D* are defined as before and *H* is a new (intermediate) hidden state. For the analysis provided here, the state *H* is selected to be *hospital admission*. This choice was carried out based on consultation with the epidemiologists on the team. It is noted that the observed transition from infected to hospitalized is well described by an exponential distribution (see Figs. 3b,3c).

Based on the fact that *I* is the initial state while *R* and *D* are both terminal states, Fig. 4b depicts a Markov chain with all possible connections including a single hidden state *H*. Our objective is to identify the transition rates 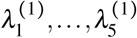 from the COVID-19 data summarized in Tables III, IV. For this purpose, it is first useful to introduce the following observed random variables:

**TABLE III:** Empirical estimates of probabilities. The quantity P(*D* |*I*) represents the case fatality rate whereas P *H*|*I* represents the fraction of cases hospitalized. The values are obtained from CDC [20].

| Quantity | Empirical Value |  |  |  | Dataset |
| --- | --- | --- | --- | --- | --- |
|  | Child | Adult | Elderly | Overall |  |
| $P(D \vert I)$ | 0.00061 | 0.014 | 0.252 | 0.029 | CDC |
| $P(H \vert I)$ | 0.0311 | 0.115 | 0.431 | 0.246 | CDC |

**TABLE IV:** Empirical estimates of expectations. The quantities E(*T* _*I*→*D*_|{*T*_*I*→*D*_< ∞}), E(*T*_*I*→*H*_ | {*T*_*I*→*H*_< ∞}), E(*T*_*I*→*R*_ {*T*_*I*→*R*_ < ∞}) represent the means of the *I* → *D, I* → *H*, and *I* → *R* datasets, respectively. Note the data in Table II have been preprocessed to remove individuals with non-positive delay values. The values come from CDC [20], NIH [21], China [27], Korea [26], Int’l [22] datasets.

| Quantity | Emp. Value | Dataset (# Datapoints) |
| --- | --- | --- |
| $E(T_{I \rightarrow D} \vert \{T_{I \rightarrow D} < \infty\})$ | 11.15 | CDC (35816) |
| $E(T_{I \rightarrow H} \vert \{T_{I \rightarrow H} < \infty\})$ | 3.82 | NIH (371) |
| $E(T_{I \rightarrow R} \vert \{T_{I \rightarrow R} < \infty\})$ | 16.46 | China, Korea, Int'l (106) |

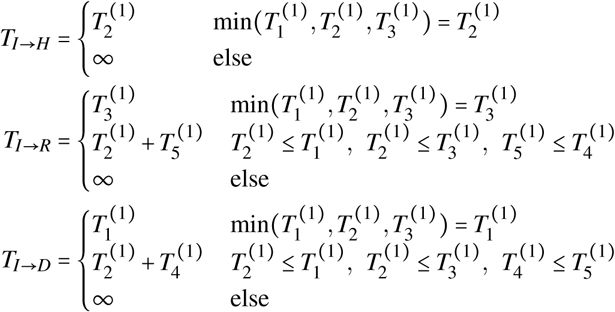

In the rest of this paper the notation for some conditional expectations is simplified. Specifically, we use E(*T*_*I*→*H*_), E(*T*_*I*→*R*_), and E(*T*_*I*→*D*_) to denote E(*T*_*I*→*H*_ |{*T*_*I*→*H*_ < ∞}), E(*T*_*I*→*R*_ |{*T*_*I*→*R*_ < ∞}), and E(*T*_*I*→*D*_ {*T*_*I*→*D*_ < ∞}), respectively.

For these random variables, the expectations are expressed as follows:

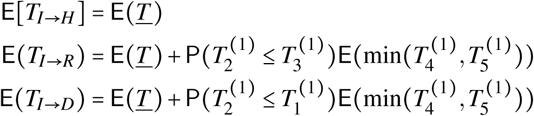

Where 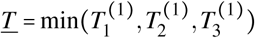. The terms on the righthand side are easily computed based on the following elementary proposition.

#### Proposition 2.

*Suppose T*_1_ ∼ exp(*λ*_1_), *T*_2_ ∼ exp(*λ*_2_)… *T*_*n*_ ∼ exp(*λ*_*n*_) *are independent exponential random variables and <u>T</u> =* min *T, T* … *T*. *Then*,

i. *<u>T</u>* ∼ exp(*λ*_1_ + *λ*_2_ + … + *λ*_*n*_
ii. 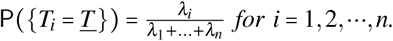

Prop. 2 yields the following equation for the three expectations:

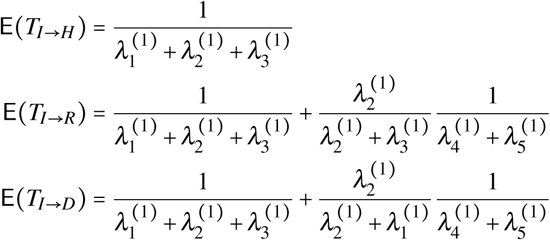

and the following equations for the two conditional probabilities:

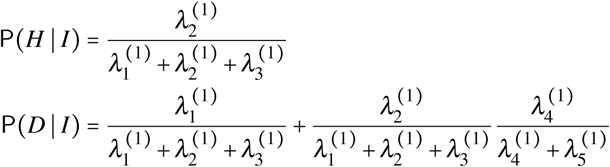

The above expectations and probabilities are readily approximated using their empirical estimates obtained from the available data (see Table II). In principle, one can then solve the five equations for five rates. If no non-negative solution exists, then the model is invalidated. Alternatively, one can derive certain necessary conditions in the form of inequalities. Prop. 3 provides one such inequality.

#### Proposition 3.

*Consider the Markov chain model depicted in Fig. 4b. Let*

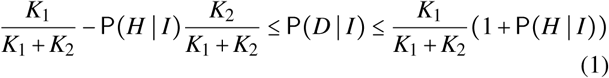

*Then*,

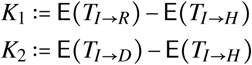

*Proof*. See the Appendix.

▪

The two inequalities are readily visualized in the (P(*D* | *I*), P(*H* | *I*)) diagram depicted in Fig. 5: the yellow region is where the inequalities are violated. By plotting the observed empirical values in the diagram, it can be seen that the model with a single hidden state is invalidated. Given that Fig. 4b represents the most general Markov chain, this also serves to show that no model with a single hidden state can fit the available data. Next, a model with two hidden states is considered.

**Fig. 5:**
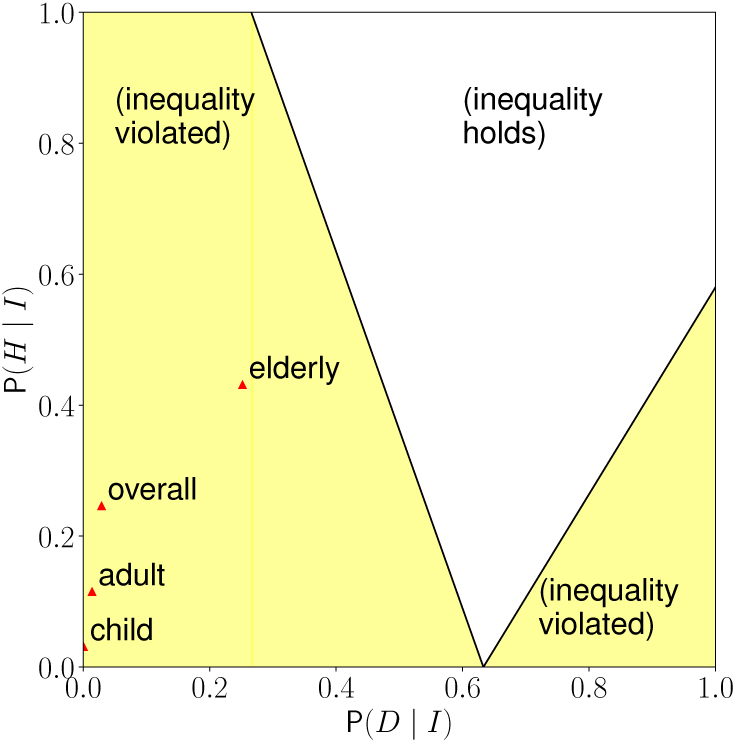
Visualization of the pair of inequalities in 1 on the (P(*D* | *I*), P(*H* | *I*)) plane. The empirical values for three age categories as well as the overall empirical value are also indicated.

### C. Two Hidden States

In Fig. 6, a Markov chain model with two hidden states is depicted. The states are denoted by {*I, H,V, R, D*}, where *H* is interpreted as before and *V* is an additional intermediate hidden state. In epidemiological terms, the state *V* has the meaning of the *end of infectious viral shedding*. The model depicted in Fig. 6 is not the most general type of model with two hidden states. For example, here it is assumed that direct *I* → *D* transition is not present. However, as is shown next, this model, apart from being epidemiologically meaningful, provides a reasonable fit to the observed data.

**Fig. 6:**
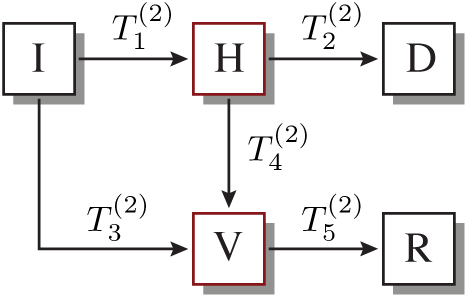
The two hidden states model, 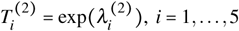.

For the model depicted in Fig. 6, the observed set of random variables are defined as follows:

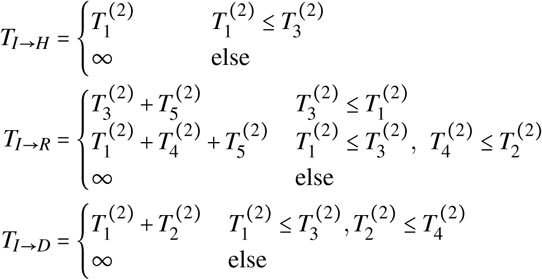

In terms of model parameters, the expectations are expressed as

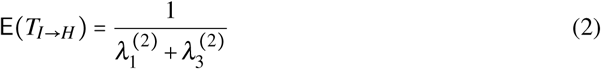

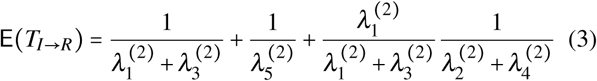

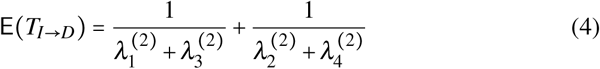

and

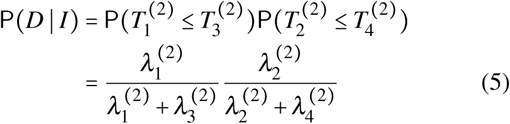

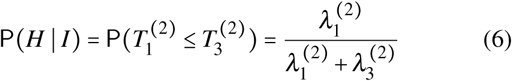

By introducing a linear parametrization, necessary and sufficient conditions for existence and uniqueness of non-negative rates are easily obtained. In this case, explicit formulae for the rates are also obtained. The result is summarized in the proposition below.

#### Proposition 4.

*Consider the Markov chain with two hidden states as depicted in Fig. 6. Provided the expressions are well-defined, the unique solution is given by*

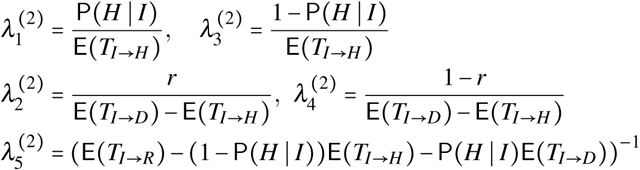

*where* 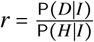. *The rates are non-negative if and only if*

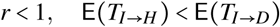

*and* 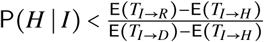.

Using these formulae, the identified rates for the three age groups are tabulated in Table V. The model is also used to obtain the distributions for the three random variables. Its comparison with the empirical distributions is depicted in Fig. 7. The expectation E(*T*_*I*→*R*_) is fitted with 5% tolerance, all other data are fitted with > 0.999% accuracy. It can be seen that the distributions for *I* → *H* and *I* → *D* are well approximated. However, the distribution fit for *I* → *R* is not as good. One explanation for the relatively poor fit for *I* → *R* could be underparametrization. Specifically, one can use (2),(4),(5) and (6) to determine 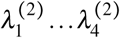 without utilizing the information of E(*T*_*I*→*R*_). Therefore, there is only one remaining degree of freedom 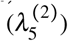 when fitting *I* → *R* distribution.

**TABLE V:** Estimated rate parameters for the two hidden state Markov chain model of Fig. 6.

| Parameter | Estimated Rate |  |  |
| --- | --- | --- | --- |
|  | Child | Adult | Elder |
| $\lambda_1^{(2)}$ | 0.009 | 0.034 | 0.090 |
| $\lambda_2^{(2)}$ | 0.003 | 0.015 | 0.095 |
| $\lambda_3^{(2)}$ | 0.285 | 0.260 | 0.119 |
| $\lambda_4^{(2)}$ | 0.155 | 0.109 | 0.067 |
| $\lambda_5^{(2)}$ | 0.068 | 0.072 | 0.158 |

**Fig. 7:**
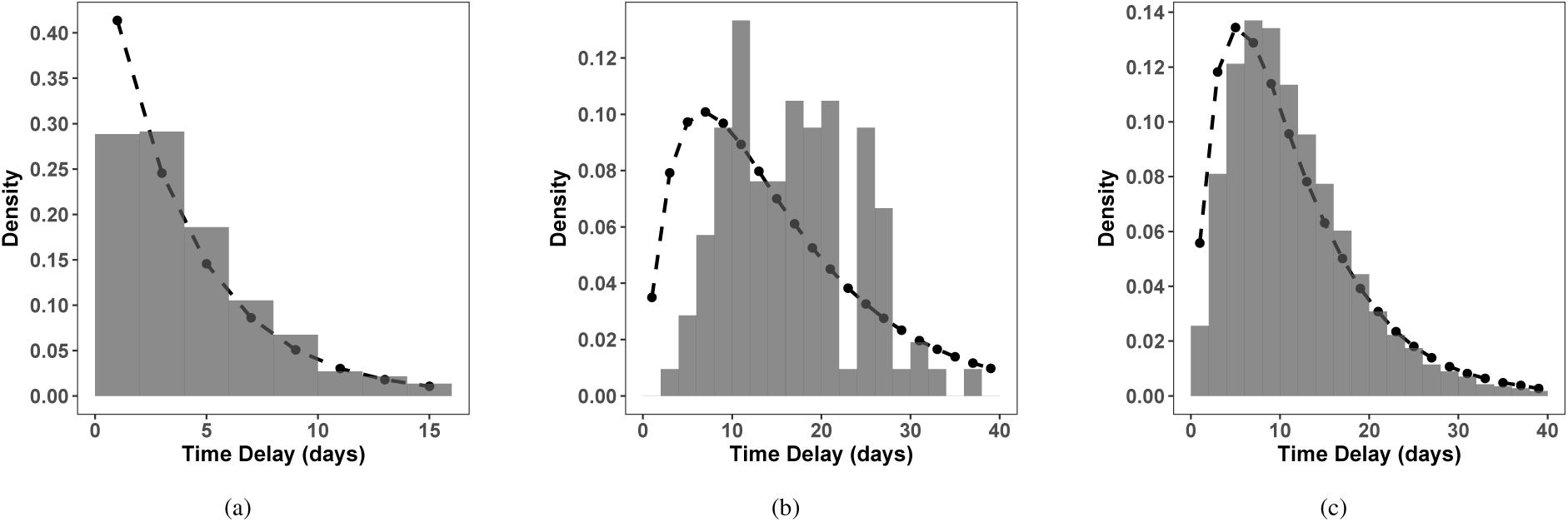
Distribution fits for (a) *I* → *H*, (b) *I* → *R*, and (c) *I* → *D* datasets based on the two hidden state model illustrated in Fig. 6.

## IV. Epidemiological Insights and Implications for Control

The analyses of Sec. III generated several interesting epidemiological insights. The analysis of the two hidden state Markov chain model, consisting of hospitalization (*H*) and end of infectious viral shedding (*V*), shows that these additional states are important to the disease progression. Seeking medical care has a dramatic effect on outcomes, and is also an indication of disease severity, with those experiencing more severe symptoms being more likely to go to the hospital. Hospitalization also is a form of quarantine, which plays a role in the chain of transmission. The end of infectious viral shedding is another key parameter in transmission and is a biologically significant event in the course of a patient’s illness. Inclusion of these two parameters suggests that these models can be successfully informed by epidemiology, and paves the way for future analyses.

Other epidemiological insights involve the differences in outcome distributions between age categories. An interesting observation gained from examining Figs. 3h and 3i is that in fatal infections, the time to death is similar among adults and the elderly since the distributions for both age categories are almost identical. This suggests that another factor, possibly comorbidity, is the driving factor behind time to death for individuals over 20 years old. In the future, it may be beneficial to stratify the data by the presence or absence of a comorbidity and build this factor into the model. Another conclusion from the *I* → *D* graph in Figs. 3g is that when children die, they die quickly. This could be due to the children having comorbidities, incorrect estimations of the time of symptom onset, or some characteristic of the pathogen that affects children differently. However, there is very limited data on children and COVID-19 in general, and thus interpretations of the data should be made cautiously. An additional observation from Figs. 3e and 3f is that adults take longer to recover than the elderly. It is important to note that the dataset for *I* → *R* is much more sparse than *I* →*D*, so inferences must be made with hesitation. This could point to different pathophysiology of the disease in different age groups, but no definitive conclusions can be drawn from these data.

While the analytical procedure outlined in this paper has provided some useful insights into the nature of COVID-19, the general approach is not confined to analysis of the current pandemic. The insights obtained from our analyses are useful for general epidemic processes, especially in the design of mitigation and control strategies. For instance, understanding the delay distributions between states is important for defining allowable response times for mitigation to be effective. This is particularly important when allocating limited resources (such as testing kits or vaccinations) among multiple regions. Looking forward, as more data are generated and collected, information regarding heterogeneity in the delay distributions will further guide response among multiple regions (especially when coupled with the growing amount of mobility data becoming available).

As a whole, this work contributes toward development of a novel strategy for building stochastic compartmental models based on Markovian principles and epidemiological data. Future data acquisition will further strengthen this methodology and allow for more insights into the disease dynamics of COVID-19.

## Data Availability

The data referred in this study are collected from publicly available resources.
Centers for Disease Control and Prevention, COVID-19 Case Surveillance Public Use Data, https://data.cdc.gov/Case-Surveillance/COVID-19-Case-Surveillance-Public-Use-Data/vbim-akqf/data, 2020.
M. Kraemer, Epidemiological data from the nCoV-2019
outbreak: Early descriptions from publicly available data, https://virological.org/t/epidemiological-data-from-the-ncov-2019-outbreak-early-descriptions-from-publicly-available-data/337, 2020.
midas network, COVID-19, https://github.com/midas-network/COVID-19/blob/master/data/cases/global/line_listings_imperial_college/international_cases_2020_08_02.csv, 2020.
Y. Xu, COVID19 inpatient cases data, https://doi.org/10.6084/m9.figshare.12195735.v3, 2020.
ThisIsIsaac,Data-Science-for-COVID-19, https://github.com/ThisIsIsaac/Data-Science-for-COVID-19/blob/master/Covid19_Dataset/patients.csv, 2020.
mrc ide,COVID19_CFR_submission,https://github.com/
mrc-ide/COVID19_CFR_submission/blob/master/data/deaths_integrated_with_linelist_17feb.csv, 2020.
Public line list and summaries of the COVID-19 outbreak in South Korea, https://github.com/parksw3/COVID19-Korea/blob/master/COVID19-Korea-2020-04-06.xlsx, 2020.
Novel Coronavirus 2019 time series data on cases, https://github.com/datasets/covid-19, 2020.
I. Dorigatti, L. Okell, A. Cori, N. Imai, M. Baguelin, S. Bhatia, A. Boonyasiri, Z. Cucunuba, G. Cuomo-Dannenburg, R. FitzJohn et al., Report 4: Severity of 2019-novel coronavirus (nCoV), Imperial College London, London, 2020.

## Appendix

*Proof of Prop. 3*. Conditional probabilities are

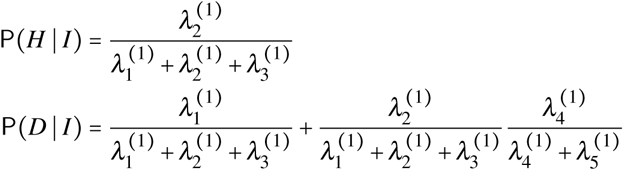

Expressing *K*_1_ and *K*_2_ in terms of parameters yields

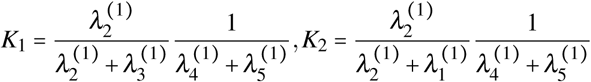

Then,

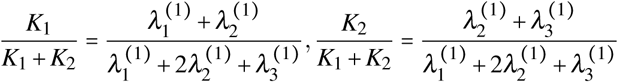

The lower bound can be shown as

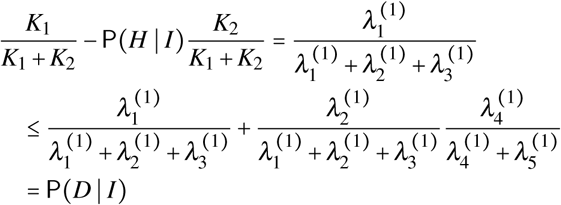

and the upper bound is given by

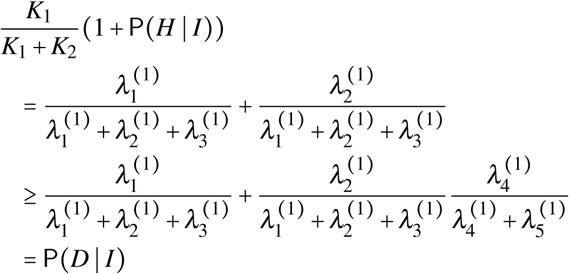

□

## Footnotes

1 Although this is a useful approximation, the transitions *I* →*R* and *I* →*D* may also be affected by the population, e.g., if the health care system is strained as a result of a large number of infected agents.

2 Anonymized line-list data consist of information on start and end date of certain epidemiological stages at an individual level.

## Notes

### Competing Interest Statement

The authors have declared no competing interest.

### Funding Statement

Research supported in part by the C3.ai Digital Transformation Institute sponsored by C3.ai Inc. and the Microsoft Corporation, in part by the Jump ARCHES endowment through the Health Care Engineering Systems Center of the University of Illinois at Urbana-Champaign, and in part by the National Science Foundation grant NSF-ECCS 20-32321.

### Author Declarations

Only the publicly available data were used in this study. Therefore, an IRB review was not required.

### Summary of Updates

Added ORCID for coauthor Prashant Mehta

